# Knowledge, attitude, and practice towards lymphatic filariasis among inhabitants of an endemic town in Oyo State, Nigeria

**DOI:** 10.1101/2022.05.30.22275754

**Authors:** Taiwo Mofadeke Jaiyeola, Ekerette Emmanuel Udoh, Abiola Basirat Adebambo

**Affiliations:** Lead City University Ibadan; Society for Family Health Abuja, Nigeria

**Keywords:** Lymphatic filariasis, knowledge, attitude, practice, mass drug administration, neglected tropical diseases, Nigeria

## Abstract

**Introduction:** The burden of lymphatic Filariasis (LF) popularly called *Ina orun* in southwest Nigeria is of serious concern and it calls for urgent attention. The study aimed at assessing the knowledge attitude and practice of inhabitants of two communities endemic with LF in Ibadan south-west Local Government Area (IBSWLGA) of Oyo State, Nigeria.

**Materials and methods:** A cross-sectional study was carried out using a semi-structured questionnaire.

**Results:** Out of 243 participants comprising of both male and female with a mean age of 35.01± 8.53 years, 35% reported ever hearing about LF. The majority (73.3%) of the participants had an overall poor knowledge of the disease, while only 26.7% have good knowledge of the disease. 26.3% of the participants knew the main cause of LF while 74.9% did not know the disease is communicable. About half (50.6%) and 42.4% of the participants did not know the symptoms and the prevention practices respectively. Only 27.2% knew that mosquitoes play a major role in LF transmission. 85.6% of the respondents were not aware of the Mass Drug Administration (MDA) program in the study area while 36.6% of participants did not know the role of MDA in preventing and controlling LF.

**Conclusion:** These results indicate poor community knowledge, inadequate prevention practices, and a lack of awareness of available programmatic efforts that the population can benefit from to tackle continuous transmission of the lymphatic filariasis disease. In conclusion, with the present low status on the disease awareness and response, increased sensitization and community interventions on LF are necessary and this calls for urgent attention.

## 1. Introduction

Lymphatic filariasis (LF), commonly called ‘elephantiasis’ is one of the most debilitating neglected diseases. It is a filarial infection caused by nematode worms *Wuchereria bancrofti, Brugia malayi*, or *timori*. The disease is transmitted to humans through the bite of infected mosquito species of the genera *aedes, anopheles, culex*, and *mansoni* (1). The infection can be acquired in childhood and can cause hidden damage to the lymphatic system. LF is currently of great public health concern in many countries of the world with up to eighty-three countries and territories being endemic, and 39 of these countries are in Sub-Saharan Africa. Nigeria has the highest number of people at risk of the infection globally followed by the Democratic Republic of Congo, Tanzania, Ethiopia and Kenya (2).

Up to 1.3 billion people are estimated to be at risk of developing the disease (3). About 68 million people are infected globally with 36 million being microfilariae carriers and 40 million symptomatic (4). Over 40 million people have been debilitated and disfigured by the disease with up to 90% being infected with *Wuchereria bancrofti* and the remainder with *Brugia malayi* or *Brugia timori* (1). Furthermore, an estimated 946 million people who live in parts of southeast Asia and sub-Saharan Africa where mosquito-borne filariasis is endemic are therefore at risk of the infection (5). After malaria, LF ranks in the list of vector-borne parasitic diseases (6). LF was recognized as the second leading cause of permanent disability. Nigeria ranks third in the list of countries endemic with LF in the world with an estimate of 22 million cases and 80 million people at risk of the disease (7). The disease has also been linked to huge economic losses affecting up to 88% of economic activity as a result of the disability linked to hydrocele in men. In addition, LF also has severe negative impact on the psychological, economic and social life of the affected population (8,9). Symptoms of LF range from acute such as filarial fever presenting as recurring pain attacks and vomiting to more chronic symptoms such as lymphodema that could progress to gross enlargement of the limbs (popularly called elephantiasis), chronic swellings on the scrotum otherwise called hydrocele (10).

In year 2000, WHO launched the global programme to eliminate lymphatic filariasis (GPELF) with the aim to interrupt transmission of lymphatic filariasis by 2020. To achieve this elimination goal, WHO recommends that a minimum of 65% population coverage should be targeted for annual mass drug administration (4). Between year 2000 and 2012, GPLEF provided more than 4.5 billion treatments in 59 countries thereby reducing the global prevalence of LF by 59%, microfilaremia prevalence by 68%, hydrocele prevalence by 50%, and lymphoedema prevalence by 25% (5). In spite of this progress, 16 countries have not yet attained the recommended coverage and ten have not yet initiated MDA. This implies that around half of the countries affected are yet to implement the recommendations of the GPELF (4).

In Nigeria, a study carried out in Ilorin, Kwara state showed that the majority (82%) of the study participants had no awareness about LF and the true cause of the infection. The respondents of the survey reported a fair understanding of LF prevention and management (8). Another study done in Kano state, Nigeria reported poor knowledge of LF symptoms and transmission. In this study, only 12% of the respondents knew that mosquito can spread LF (11).

Assessing the knowledge, attitude and practices of citizens in endemic communities concerning the disease and its elimination is important to evaluate the control effort being carried out and provide information on the potential risks these communities remain exposed to. More so, data on awareness and response of endemic communities is useful for programming to reduce vector borne disease burden, as well as help in developing and implementing effective and sustainable control program for the disease. In addition, recognizing a community’s participation in tackling the disease is very critical to the success of any elimination program. Therefore, this present study was carried out to assess the knowledge, attitude and practice of the community residents about LF with regards to the causes, transmission, prevention and control practices.

## 2. Materials and Methods

### Study settings

Ibadan South-west LGA is in Ibadan metropolis and is one of the 33 LGAs in the state. The LGA has an estimated population of 190,672 inhabitants (12). LF is endemic in the LGA with prevalence of 21% from the LGA surveillance records. The disease is co-endemic with onchocerciasis in this LGA. This study was cross-sectional in design. Total sample size was 243 and interviews were conducted for this number of participants. A multistage sampling technique was used to sample for the participants. Firstly, two wards were purposively selected out of the twelve wards in the LGA. These two wards were selected because of established cases of the disease in the wards from records in the disease surveillance office of the IBSWLGA. In the next stage, one community each was selected from each of the two wards. Finally, a systematic random sampling process was used to sample households in each of the selected communities. Respondents who met inclusion criteria in households selected were interviewed. The study population included males and females aged 18 years and above who had been resident in the two study communities for at least one year, and gave a written consent to participate in the study.

### Data collection and analysis

A semi-structured questionnaire was developed and pretested in a different LF endemic area to ensure consistency, reliability, and appropriateness of language before the questionnaire was deployed for data collection in the study areas. The questionnaire was translated from English into the local language Yoruba for interviews with participants who did not understand English. Information from respondents captured with the questionnaire were socio-demographic characteristics, knowledge about LF including causes, signs and symptoms, mode of transmission, prevention and management of LF. Also, information on attitude and practices of respondents towards prevention and control of the disease, and participation in Mass Drug Administration (MDA) program was assessed. Data collection was done by trained interviewers who administered the questionnaire electronically using Kobocollect application on android devices.

Data was exported from Kobocollect into Statistical package for social sciences version 20. Data was cleaned and analyzed. Questions on knowledge relating to causes of LF, mode of transmission, prevention, and knowledge of control program through MDA were summed into a composite knowledge-score variable. Respondents whose total scores were less than the mean of the composite knowledge-score variable were classified as having poor knowledge, while respondents whose total scores were equal to or greater than the mean knowledge score were classified as having good knowledge. Questions related to practice towards LF and its elimination were also summed into a composite practice-score variable. Respondents whose scores were less than the mean score were grouped as having poor practice towards LF, while those whose scores were equal to or greater than the mean were considered as having good practice towards LF. All results are presented using descriptive statistics using frequency and percentages.

## 3. Results

### 3.1. Sociodemographic characteristics of respondents

Mean age of respondents was 35.01 ± 8.53 years with males making 50.6% and female 49.4% of the sample population. More than half of the respondents were married (58.4%), while 30.9% were single. Majority were in skilled occupation (43.2%) and 32.9% were in unskilled occupation. Concerning the educational level of participants, about 40% reported to have completed secondary education, 17.3% had a tertiary education and 3.3% reported no education. About half of the respondents were Christians (49.8%) and 41.6% were Muslims, while 1.2% did not report having a religious affiliation. Majority of the respondents (60.1%) had resided in the study communities for more than ten years, 21% had been resident in the community for about 5-10 years, only a few (7.8%) had spent about a year in the study area. Table 1 shows the socio-demographic profile of the respondents.

**Table 1.** Socio-demographic characteristics of respondents.

| Variable | Frequency<br>N = 243 | % |
| --- | --- | --- |
| Age group |  |  |
| 20-29 | 84 | 34.6 |
| 30-39 | 99 | 40.7 |
| 40-49 | 38 | 15.6 |
| 50-59 | 22 | 9.1 |
| **Sex** |  |  |
| Males | 123 | 50.6 |
| Females | 120 | 49.4 |
| **Marital status** |  |  |
| Single | 75 | 30.9 |
| Married/ Cohabiting | 146 | 60.1 |
| Divorced, separated /widowed | 20 | 8.2 |
| No Response | 2 | 0.8 |
| **Occupation** |  |  |
| Professionals | 3 | 1.2 |
| Highly Skilled | 32 | 13.2 |
| Skilled | 105 | 43.2 |
| Semi-skilled | 17 | 7.0 |
| Unskilled | 80 | 32.9 |
| Unemployed | 6 | 2.4 |
| None | 8 | 3.3 |
| **Education** |  |  |
| No education | 8 | 3.3 |
| Some primary | 3 | 1.2 |
| Completed primary | 21 | 8.6 |
| Some secondary | 72 | 29.6 |
| Completed secondary | 97 | 39.9 |
| Tertiary | 42 | 17.3 |
| **Religion** |  |  |
| Christianity | 121 | 49.8 |
| Islam | 101 | 41.6 |
| Others | 13 | 5.3 |
| No response | 8 | 3.2 |
| **Length of stay in**<br>**Community (years)** |  |  |
| About 1 | 19 | 7.8 |
| >1-5 | 27 | 21.0 |
| >5-10 | 51 | 21.0 |
| >10 | 146 | 60.1 |

### 3.2 Knowledge of lymphatic filariasis among respondents

Our result showed that majority of participants had an overall poor knowledge of LF (73.3%), with less than a third showing good knowledge of LF from the analysis (26.7%) (Table 2). Only 35% of the study respondents had heard about the disease. Regarding respondents’ knowledge on the symptoms of LF, only about one-third (33.3%) knew that LF may not show any sign, more than a third of the respondents correctly associated the disease with lymphedema (33.3%), elephantiasis (33.7%), hydrocele (30%) and about half (50.6%) of the study participants did not know any symptom of LF.

**Table 2.**
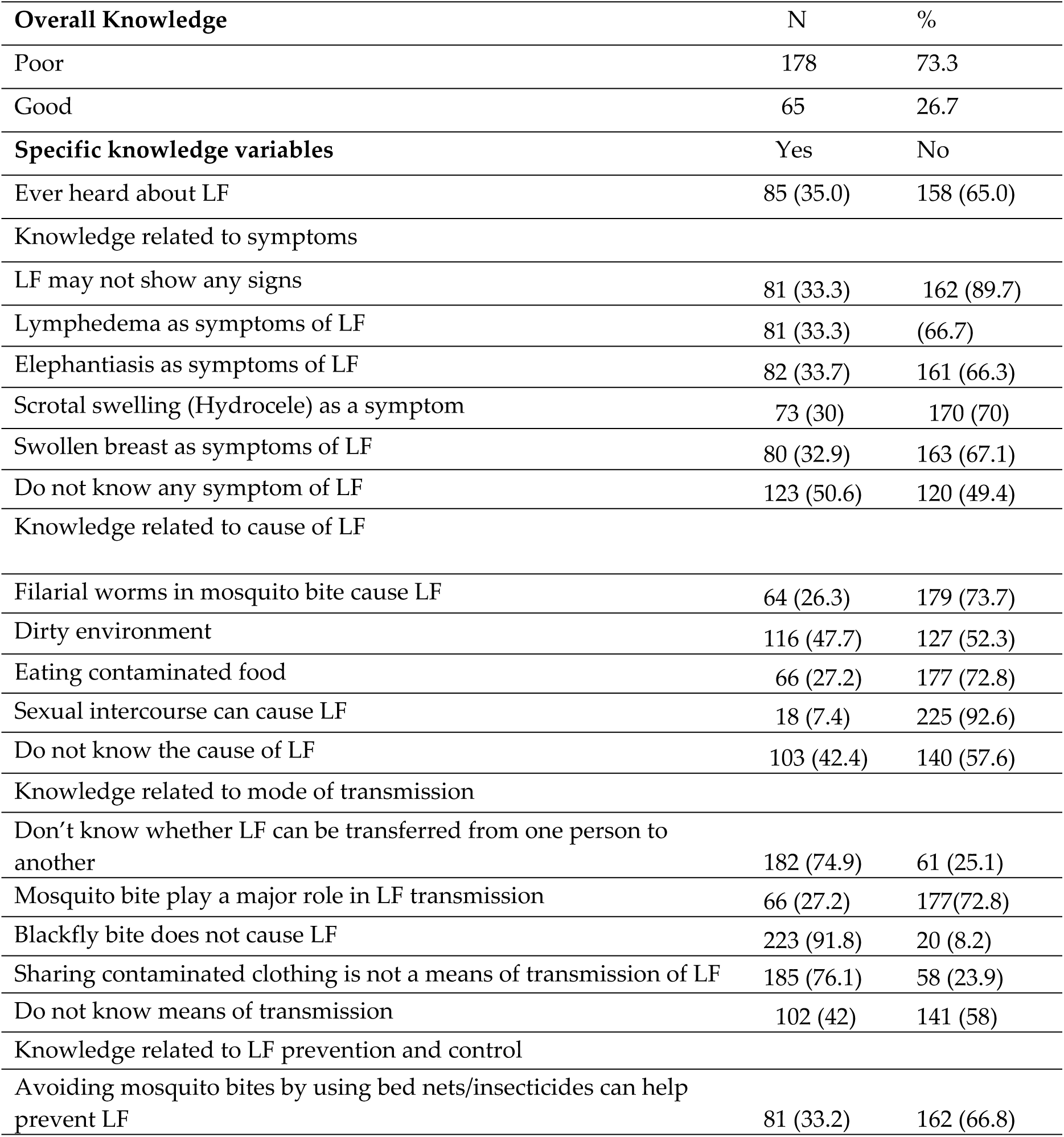
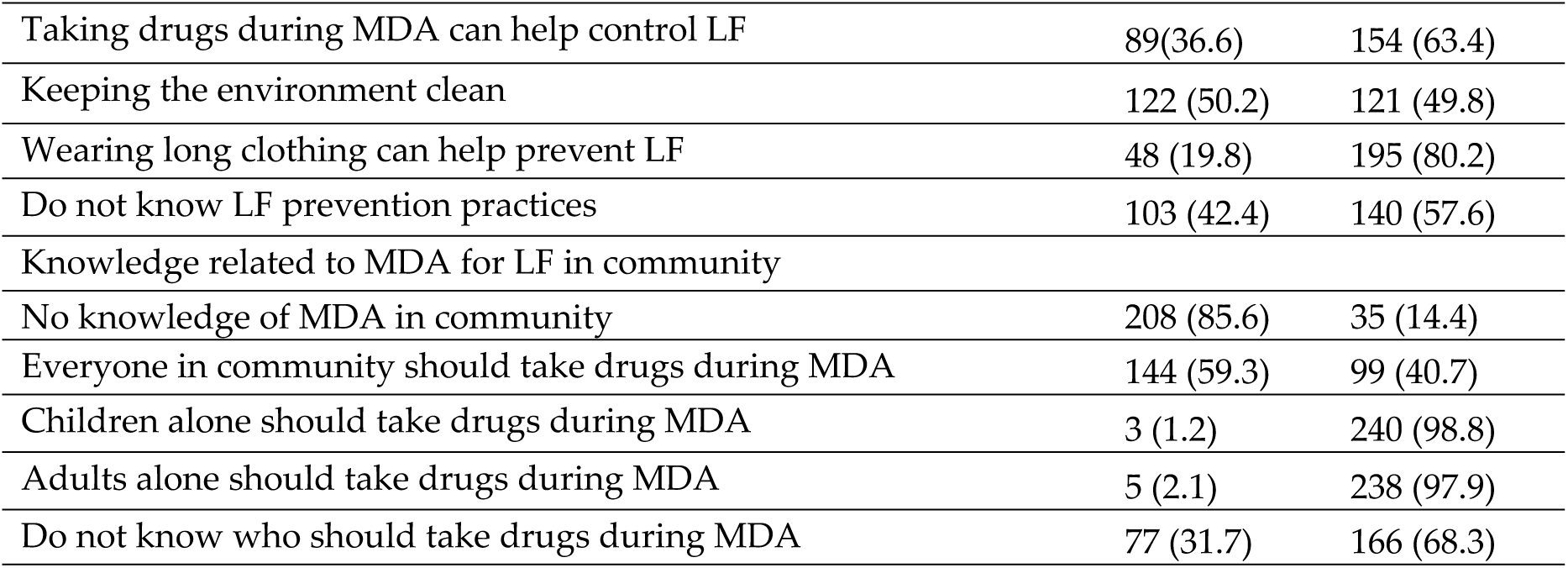
Knowledge of Lymphatic filariasis among respondents.

Only 26.3% of respondents knew that filarial worm from mosquito bites is the cause of LF. Meanwhile, other reasons like dirty environment (47.7%) and eating contaminated food (27.2%) were wrongfully given as the cause of LF, with up to 42.4% unaware of what the cause of LF was. Most of the respondents did not know whether LF can be transferred from one person to another (74.9%). Some respondents knew that mosquitoes play a major role in the transmission of the disease (27.2%). The participants also reported dirty environment (47.7%), eating contaminated food (35%), and sharing contaminated clothing (23.9%) as some of the modes of transmission of LF. Some respondents also attributed transmission of LF to blackfly bite (8.2%). The percentage that reported not knowing the mode of transmission of LF was 42%.

About on third (33.2%) of participants knew that LF could be prevented by avoiding mosquito bites, with 36.6% reporting that drugs for LF would help to control LF. Other reported measures such as keeping the environment clean (50.2%) wearing long clothing (19.8%), and using bed nets or insecticide (43.6%) were reported for LF prevention. Most of the respondents did not know about the MDA program (85.6%) for the control of LF in the community. Over half of the respondents mentioned that everyone in the community should take drugs during MDA (59.3%) even though almost one-third do not know who should take drugs during MDA (31.7%).

### 3.3 Attitude towards lymphatic filariasis and its elimination

Attitude towards lymphatic filariasis was assessed and is presented in Table 3. Most of the respondents reported they would be ashamed about the disease if they or a relation became sick from LF (53.5%). The percentage who reported that they would be sad/worried if they or their family members becomes sick from LF was 44.9%. Majority (69.1%) of the respondents think the disease needs treatment and treatment options reported include: use of modern medicines (19.8%), use of traditional medicines (5.3%) and use of both modern and traditional (74.9%) medicines.

**Table 3:**
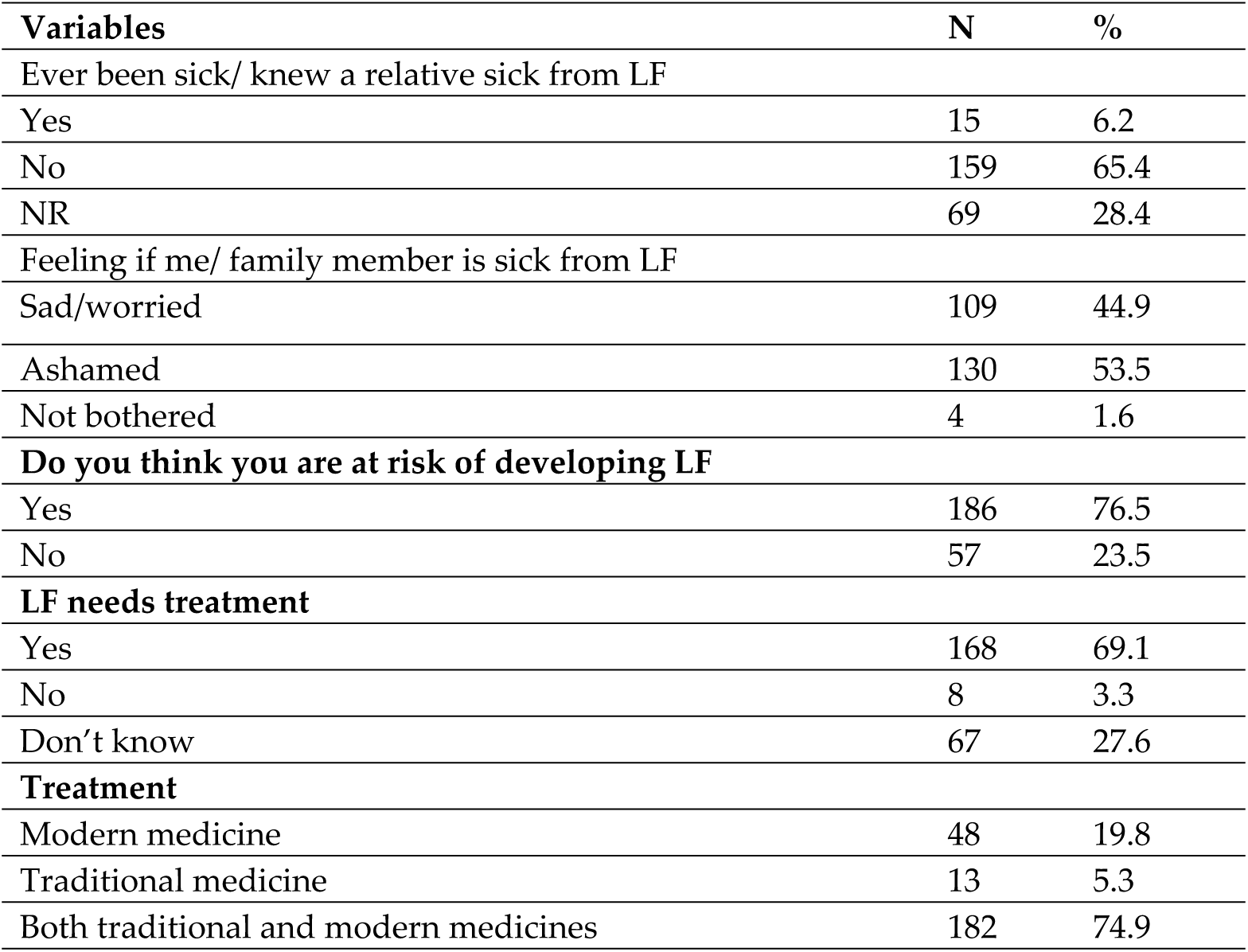
Respondents’ Attitude towards LF and its Control.

| **Variables** | **N** | **%** |
| --- | --- | --- |
| Ever been sick/ knew a relative sick from LF |  |  |
| Yes | 15 | 6.2 |
| No | 159 | 65.4 |
| NR | 69 | 28.4 |
| Feeling if me/ family member is sick from LF |  |  |
| Sad/worried | 109 | 44.9 |
| Ashamed | 130 | 53.5 |
| Not bothered | 4 | 1.6 |
| **Do you think you are at risk of developing LF** |  |  |
| Yes | 186 | 76.5 |
| No | 57 | 23.5 |
| **LF needs treatment** |  |  |
| Yes | 168 | 69.1 |
| No | 8 | 3.3 |
| Don't know | 67 | 27.6 |
| **Treatment** |  |  |
| Modern medicine | 48 | 19.8 |
| Traditional medicine | 13 | 5.3 |
| Both traditional and modern medicines | 182 | 74.9 |

### 3.4 Practice towards lymphatic filariasis and its elimination

Practice towards LF control and elimination are presented in Table 4. Less than 10% of the respondents took drugs at the last round of MDA for LF. Majority of the respondents didn’t know about the MDA program for LF. Other reasons given for not taking drugs include: I don’t think I’m at risk of LF (6.6%), taking other precautions (3.3%), no reason given (24.3%). Activities undertaken by respondents to prevent LF reported includes: avoiding mosquito bites (44%), keeping clean environment (23.9%) and about 16% of the study participants don’t do anything to prevent LF. Even though, only a few of this study respondents reported being affected with the disease (6.2%), more than half of this affected few visited traditional homes for treatment. Practice towards LF and its control among the respondents was generally poor with only 21.8% having a good practice score.

**Table.** Respondents’ practice towards LF and its Control.

**Table Respondents' practice towards LF and its Control**
| Variables | N | % |
| --- | --- | --- |
| **Received drugs at last round** |  |  |
| Yes | 20 | 8.4 |
| No | 223 | 91.8 |
| **Why drug was not received** |  |  |
| Didn't know about MDA | 157 | 64.6 |
| I take other precautions | 52 | 21.4 |
| No Reason | 34 | 14.0 |
| **Other Actions taken to prevent LF** |  |  |
| Avoiding mosquito bite | 107 | 44 |
| Keeping clean environment | 58 | 23.9 |
| Don't do anything | 40 | 16.5 |
| **Place of treatment** |  |  |
| Health facility | 6 | 40 |
| Traditional healers' home | 9 | 60 |
| **Overall practice on LF disease** |  |  |
| Poor | 190 | 78.2 |
| Good | 53 | 21.8 |

## 4.0 Discussion

This study revealed that although the communities in the study area are endemic for LF, majority of the population are unaware of the disease, signifying that information and awareness creation on the disease remains very low in these communities. Awareness creation would allow the population to have specific knowledge, for example, on the causal agent and the routes of transmission of the disease. In the present study, we found that the population lacked the knowledge or had wrong knowledge of the causal agent of LF. For instance, respondents attributed the cause of LF to numerous reasons unrelated to the cause of the disease. This included reasons such as eating contaminated food, sexual intercourse, or dirty environment, rather than to the actual cause which is filarial worms in mosquito bites. Our findings on misconception about causes of LF corroborates findings from a previous study carried out in Benue state, Nigeria where majority in the study reported causes of LF as lack of personal hygiene, walking long distances, stepping on charm (13).

Our findings also showed that majority of the respondents did not know that LF is a communicable disease, which suggests that precautionary measures may not be taken against the disease by many members of the affected community thereby increasing the risk of transmission. Similar high lack of knowledge on the transmission of the LF disease is common in other endemic countries such as India, where a study reported a populations’ lack of knowledge on filariasis as a communicable disease (14). Only about 27% of respondents in our study knew that LF is transmitted by mosquito bites. Lack of knowledge of transmission agent for LF can pose a challenge to combating or implementing effective vector control measures. For example, the population would not see the need for use of mosquito nets to protect against mosquito bites in the household. It was also observed that only one-third of the study participants knew that avoiding mosquito bites can help to prevent LF (33.2%). Our findings is similar with other studies in India and Nepal where knowledge about the vector for LF is still rife, especially in the rural areas (15),(16). Another study have also reported that many LF endemic communities do not know the significance of reducing contact with mosquitos to prevent LF (17).

Lack of adequate knowledge on simple preventive measures such as use of insecticide treated nets/ insecticide to prevent mosquito bites, taking of drugs during the MDA program put the community at increased risk of the disease. About one third (36.6%) of the respondents know about the MDA to prevent LF while 42% do not know LF prevention measures. However, these findings are contrary to the findings of a study in Plateau state, Nigeria where the majority reported that MDA and vector control can be used to prevent LF (6). There is therefore the need to increase awareness and coverage of the MDA in the study area. If adequate knowledge of prevention is lacking, then control measures will be inappropriate, this will in turn make control efforts targeted at the disease ineffective (14).

Concerning respondents’ attitude towards LF, we found that although about 6.2% of the sample reported ever being sick or had relations sick from LF, most respondents felt it was important to treat. Unfortunately, the perception of being at risk of the disease amongst the sample was quite poor as 76.6% did not think they were at risk of getting the disease. This finding is similar to results from the study in Plateau state where about half of study participants did not feel they were at risk of being infected with the disease (6). Another study in Taraba state Nigeria had a similar finding where respondents did not consider themselves susceptible to the disease (18). This low level of perceived risk may be associated with the overall low knowledge of the disease in the study area. This study showed that many of the respondents did not receive drugs during the last MDA mainly because of lack of knowledge of the MDA program. This is however in contrast to a study carried out in Telangana State of India where the majority (88%) of the respondents consumed DEC tablets during MDA because majority knew about MDA in their community (14).

## 5. Conclusion

The study has shown that knowledge of LF in the communities surveyed was generally low, especially as regards the mode of transmission. Half of the study participants do not know the symptoms; a considerable proportion does not know the prevention and control measures. The role of mosquitoes in the transmission of the disease is not well understood hence not much is done by community members to prevent mosquito bites. The survey showed coverage of MDA in the study area is very low but the people also do not understand the role of MDA to prevent and control the disease. There is a need for increased awareness of the disease because a prevalence as high as 21% reported by the state MOH and the poor knowledge found among the study participants seem alarming. Awareness can be increased through health education on the causes, mode of transmission, symptoms, prevention, treatment, and control of the disease. The need for vector control and improved treatment coverage is imperative while emphasizing the goal of the MDA program to the communities.

## Data Availability

All relevant data are within the manuscript and its Supporting Information files.

## Acknowledgments

This study acknowledges the input of Dr. Olubukola Adebayo-Tosin in reviewing the manuscript to improve the outlook

## Supplemental Materials

## Authors Contributions

Conceptualization: Jaiyeola T.M.; methodology, Jaiyeola T.M and Udoh E.E.; validation, Jaiyeola T.M, Udoh E.E and Adebambo B.A; formal analysis, Jaiyeola T.M and Udoh. E.E; investigation, Adebambo B.A; resources, Jaiyeola T.M, Adebambo B.A.; data curation, Udoh E. E and Jaiyeola T.M.; writing—original draft preparation, Jaiyeola T.M.; writing— review and editing Udoh E.E and Jaiyeola T.M; visualization, Jaiyeola T.M, supervision, Udoh E.E.; project administration, Udoh E.E.; funding acquisition, Jaiyeola T.M and Adebambo B.A. All authors have read and agreed to the published version of the manuscript.

## Institutional Review Board Statement

Ethical clearance was obtained from the Research ethical review committee board of the Oyo State ministry of health AD 13/479/4436^A^. Permission was sought from the authorities of the LGA. Survey started with the informed consent being read to each participant with the option of agreeing or not agreeing to participate in the survey. Those who agreed to participate were those who proceeded with the interview. All information obtained from the respondents was kept confidential. The study was conducted in accordance with the Declaration of Helsinki, and approved by the Ethical review board of Oyo State Ministry of Health (protocol code AD 13/479/4436^A^ and 31^st^ August, 2021.

## Informed Consent Statement

Informed consent was obtained from all subjects involved in the study

## Data Availability Statement

Not applicable.

## References

1. WHO. Lymphatic filariasis [Internet]. 2021 [cited 2022 Mar 2]. Available from: https://www.who.int/news-room/fact-sheets/detail/lymphatic-filariasis

2. Hotez PJ, Kamath A. Neglected tropical diseases in sub-Saharan Africa: Review of their prevalence, distribution, and disease burden. PLoS Negl Trop Dis. 2009;3(8):2–11.

3. Lustigman S, Prichard RK, Gazzinelli A, Grant WN, Boatin BA, Mccarthy JS, et al. A Research Agenda for Helminth Diseases of Humans: The Problem of Helminthiases. PLoS Negl Trop Dis [Internet]. 2012 [cited 2021 Dec 21];6(4). Available from: http://www.cnpq.

4. Ndeffo-Mbah ML, Galvani AP. Global elimination of lymphatic filariasis. Lancet Infect Dis [Internet]. 2017;17(4):358–9. Available from: http://dx.doi.org/10.1016/S1473-3099(16)30544-8

5. Ramaiah KD, Ottesen EA. Progress and Impact of 13 Years of the Global Programme to Eliminate Lymphatic Filariasis on Reducing the Burden of Filarial Disease. PLoS Negl Trop Dis. 2014;8(11).

6. Azzuwut MP, Sambo MN, Hadejia IS. Assessment of the knowledge, attitude and practices related to the treatment and prevention of lymphatic filariasis among the adult residents of Bokkos local government area of Plateau state, Nigeria. Jos J Med. 2012;6(3):16–8.

7. World Health Organization. Global Programme to Eliminate Lymphatic Filariasis. Annual Report on Lymphatic Filariasis 2002. 2002.

8. Amaechi EC, Ohaeri CC, Ukpai OM, Nwachukwu PC, Ukoha UK. Lymphatic filariasis: knowledge, attitude and practices among inhabitants of an irrigation project community, North Central Nigeria. Asian Pacific J Trop Dis. 2016;6(9):709–13.

9. Haddix AC KA. Lymphatic filariasis: Economic aspects of the disease and programmes for its elimination. Trans R Soc Trop Med Hyg. 2000;94(6):592–593.

10. World Health Organization. Lymphatic filariasis: Reasons for Hope. In 1997.

11. Dogara M. Survey of knowledge, attitudes and perceptions (KAPs) of lymphatic filariasis patients in Kano State, Nigeria. Int Res J Public Environ Heal [Internet]. 2014;1(10):207–10. Available from: http://www.journalissues.org/IRJPEH/http://dx.doi.org/10.15739/irjpeh.010

12. Ibadan South West Local Government Area [Internet]. [cited 2021 Dec 28]. Available from: https://www.manpower.com.ng/places/lga/678/ibadan-south-west

13. Agbo OE, Ochanya OJ. Clinical epidemiology of lymphatic filariasis and community practices and perceptions amongst the Ado people of Benue State, Nigeria. Vol. 5, African Journal of Infectious Diseases. 2011. p. 47–53.

14. Medoju A, Gedam CM, Lakshman SM. Knowledge, Attitude and Practices (KAP) about Lymphatic Filariasis and Perception Regarding Socio-Economic Status of Diseased Person among Inhabitants of Erstwhile Warangal District, Telangana State. 2019 [cited 2022 Jan 6];2(4):65–73. Available from: www.ijsrm.humanjournals.comwww.ijsrm.humanjournals.com

15. Shercand J, Obsomer Valerie TGD and Hm, D. Mapping of lymphatic filariasis in Nepal. Filaria J. 2003;2(7).

16. Hopkins DR, Eigege A, Miri ES, Gontor I, Ogah G, Umaru J, et al. Lymphatic filariasis elimination and schistosomiasis control in combination with onchocerciasis control in Nigeria. Am J Trop Med Hyg. 2002;67(3):266–72.

17. Wynd S, Melrose WD, Durrheim DN, Carron J, Gyapong M. Bulletin of the World Health Organization. Bull World Health Organ. 2007;85(6):493–8.

18. Ogbonnaya LU, Okeibunor JC. Sociocultural Factors Affecting the Prevalence and Control of Lymphatic Filariasis in Lau Local Government Area, Taraba State: http://dx.doi.org/102190/AY5A-QAY4-6H8D-VEL7 [Internet]. 2016 Aug 1 [cited 2022 Jan 6];23(4):341–71. Available from: https://journals.sagepub.com/doi/abs/10.2190/AY5A-QAY4-6H8D-VEL7

